# A Web-based, Mobile Responsive Application to Screen Healthcare Workers for COVID Symptoms: Descriptive Study

**DOI:** 10.1101/2020.04.17.20069211

**Authors:** Haipeng (Mark) Zhang, Dimitar Dimitrov, Lynn Simpson, Balaji Singh, Nina Plaks, Steve Penny, Jo Charles, Rose Sheehan, Steve Flammini, Shawn Murphy, Adam Landman

## Abstract

**Background:** The COVID-19 pandemic has impacted over 1 million people across the globe, with over 330,000 cases in the United States. To help limit the spread in Massachusetts, the Department of Public Health required that all healthcare workers must be screened for symptoms daily – individuals with symptoms may not work. We rapidly created a digital COVID-19 symptom screening tool for a large, academic, integrated healthcare delivery system, Partners HealthCare, in Boston, Massachusetts.

**Objective:** We describe the design and development of the COVID-19 symptom screening application and report on aggregate usage data from the first week of use across the organization.

**Methods:** Using agile principles, we designed, tested and implemented a solution over the span of a week using progressively custom development approaches as the requirements and use case become more solidified. We developed the minimum viable product (MVP) of a mobile responsive, web-based self-service application using REDCap (Research Electronic Data Capture). For employees without access to a computer or mobile device to use the self-service application, we established a manual process where in-person, socially distanced screeners asked employees entering the site if they have symptoms and then manually recorded the responses in an Office 365 Form. A custom .NET Framework application was developed solution as COVID Pass was scaled. We collected log data from the .NET application, REDCap and Office 365 from the first week of full enterprise deployment (March 30, 2020 – April 5, 2020). Aggregate descriptive statistics including overall employee attestations by day and site, employee attestations by application method (COVID Pass automatic screening vs. manual screening), employee attestations by time of day, and percentage of employees reporting COVID-19 symptoms

**Results:** We rapidly created the MVP and gradually deployed it across the hospitals in our organization. By the end of the first week of enterprise deployment, the screening application was being used by over 25,000 employees each weekday. Over the first full week of deployment, 154,730 employee attestation logs were processed across the system. Over this 7-day period, 558 (0.36%) employees reported positive symptoms. In most clinical locations, the majority of employees (∼80-90%) used the self-service application, with a smaller percentage (∼10-20%) using manual attestation. Hospital staff continued to work around the clock, but as expected, staff attestations peaked during shift changes between 7-8am, 2-3pm, 4-6pm, and 11pm-midnight.

**Conclusions:** Using rapid, agile development, we quickly created and deployed a dedicated employee attestation application that gained widespread adoption and use within our health system. Further, we have identified over 500 symptomatic employees that otherwise would have possibly come to work, potentially putting others at risk. We share the story of our implementation, lessons learned, and source code (via GitHub) for other institutions who may want to implement similar solutions.

## Introduction

To date, there are over 1.2 million confirmed cases of COVID-19 throughout the world with over 330 thousand in the United States. This number is rapidly growing and the United States has become the epicenter of COVID-19. ^1^ By April 5^th^, in the Commonwealth of Massachusetts, there have been a total of 12,500 confirmed cases of COVID-19 and 231 deaths. ^2^ The Institute for Health Metrics and Evaluation at the University of Washington projects that peak resource utilization in Massachusetts will occur in mid-April with potentially 373 COVID-19 deaths at peak and an intensive care unit bed shortage measured in the thousands. ^3^. With the exponentially rising number of cases and limited resources, there is a growing recognition that scalable solutions such as digital technology can be used to address issues that arise from pandemics such as COVID-19. ^4^

To limit the spread and “flatten the curve”, on March 16^th^, 2020, the Massachusetts Department of Public Health (MDPH) and the Commissioner of Public Health issued an order that Massachusetts hospitals must screen all visitors, including employees, for symptoms of a respiratory infection (fever, cough, shortness of breath, or sore throat) and those individuals with any symptoms should not be permitted to visit.^5^ Shortly after, our institution enacted a policy that all employees working in a patient care facility must wear a face mask while working as another measure to limit the spread of COVID-19 within the healthcare workforce. ^6^

We created a digital symptom screening and attestation tool whose output functions as the daily facility and face mask pass for the employee called COVID Pass. Using a prototype-driven innovation model combined with a transition to a more traditional custom development team as the application requirements matured allowed our group to rapidly deploy and refine a solution as it was released and approached scale. In under 2 weeks, COVID Pass transitioned from a paper proposal to enterprise supported solution that is currently being used by over 25,000 employees daily.

In this paper, we describe the design, development and use of the application and make the code available for other institutions seeking a similar solution.

## Methods

This study took place at Partners HealthCare (Partners), a not-for-profit, academic, integrated health care delivery system in Boston, Massachusetts. Partners includes Brigham and Women’s Hospital, Massachusetts General Hospital, community and specialty hospitals, a physician network, community health centers, home care, a health insurance plan, and other health related services. The largest private employer in Massachusetts, Partners has approximately 74,000 employees, including physicians, nurses, scientists, and caregivers.

Complying with the MDPH requirement for symptom screening in such a large, geographically distributed organization was going to be difficult. The Partners HealthCare Chief Human Resources Officer recognized an opportunity to link the distribution of masks (something that employees wanted) with completion of the symptom screen (something that might be more difficult to have all employees complete). Human Resources and Occupational Health provided initial requirements including an application that would enable employees who must work onsite at a facility that provides direct patient care to be able to self-screen for symptoms concerning for COVID-19 infection prior to being allowed into the facility. This application needed to be mobile-responsive, provide guidance to the employee about next steps if they do indicate symptoms are present, create a pass that would be “glance-able” to entrance way screening staff, and be able to export user logs on at least a daily basis. In addition, a solution was also needed for employees that are unable to utilize the electronic self-screening tool to be processed via a manual pathway where screeners would ask patients about symptoms and record the results.

Using agile principles, we designed, tested and implemented a solution over the span of a week using progressively custom development approaches as the requirements and use case become more solidified. Based on the requirements, we developed the minimum viable product (MVP) of the self-service application using REDCap (Research Electronic Data Capture) for the speed that we could develop a functional prototype along with the prebuilt systems to export data from this solution. REDCap is a secure, web-based software platform designed to support data capture for research studies, providing 1) an intuitive interface for validated data capture; 2) audit trails for tracking data manipulation and export procedures; 3) automated export procedures for seamless data downloads to common statistical packages; and 4) procedures for data integration and interoperability with external sources.^7,8^

The self-service application, built in REDCap, was called COVID Pass. The application required login using Partners network login user ID and password. The application then authenticated the user against active directory and used the login ID to lookup the employees first name, last name, email address, and employee ID. The employee would review this information and select whether they have symptoms of COVID from a list of symptoms determined by infection control leaders (Figure 1A). If the employee selected “no symptoms”, they were required to attest with their initials, and were then provided a pass to enter the facility for the day. The pass was displayed on the screen and a copy was automatically sent to their email address (Figure 1B). Employees indicating one or more symptoms were directed not to come to work and to call their manager and occupational health (Figure 1C). A copy of the relevant information was also sent to the employee’s email address.

**Figure 1A.**
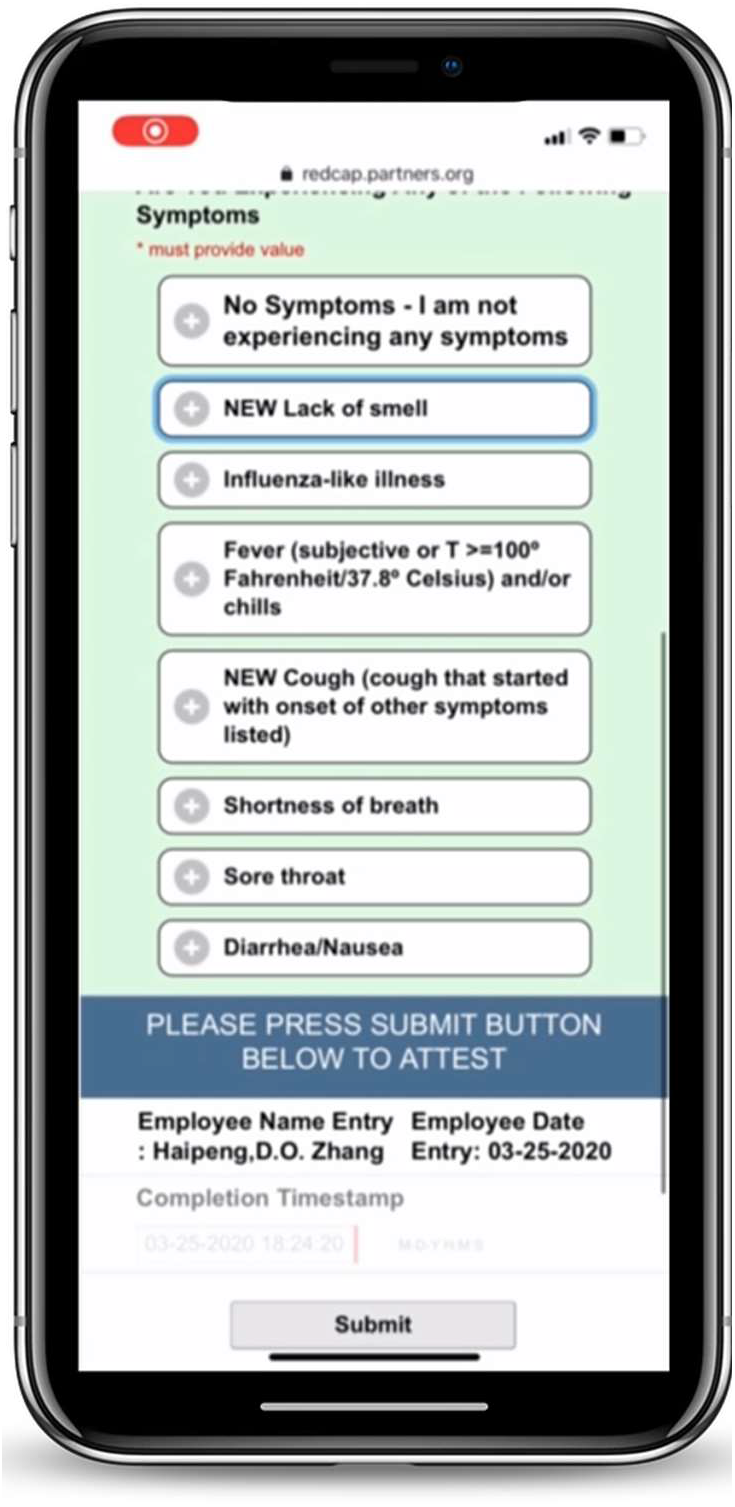
COVID Pass employee symptom reporting screen.

**Figure 1B.**
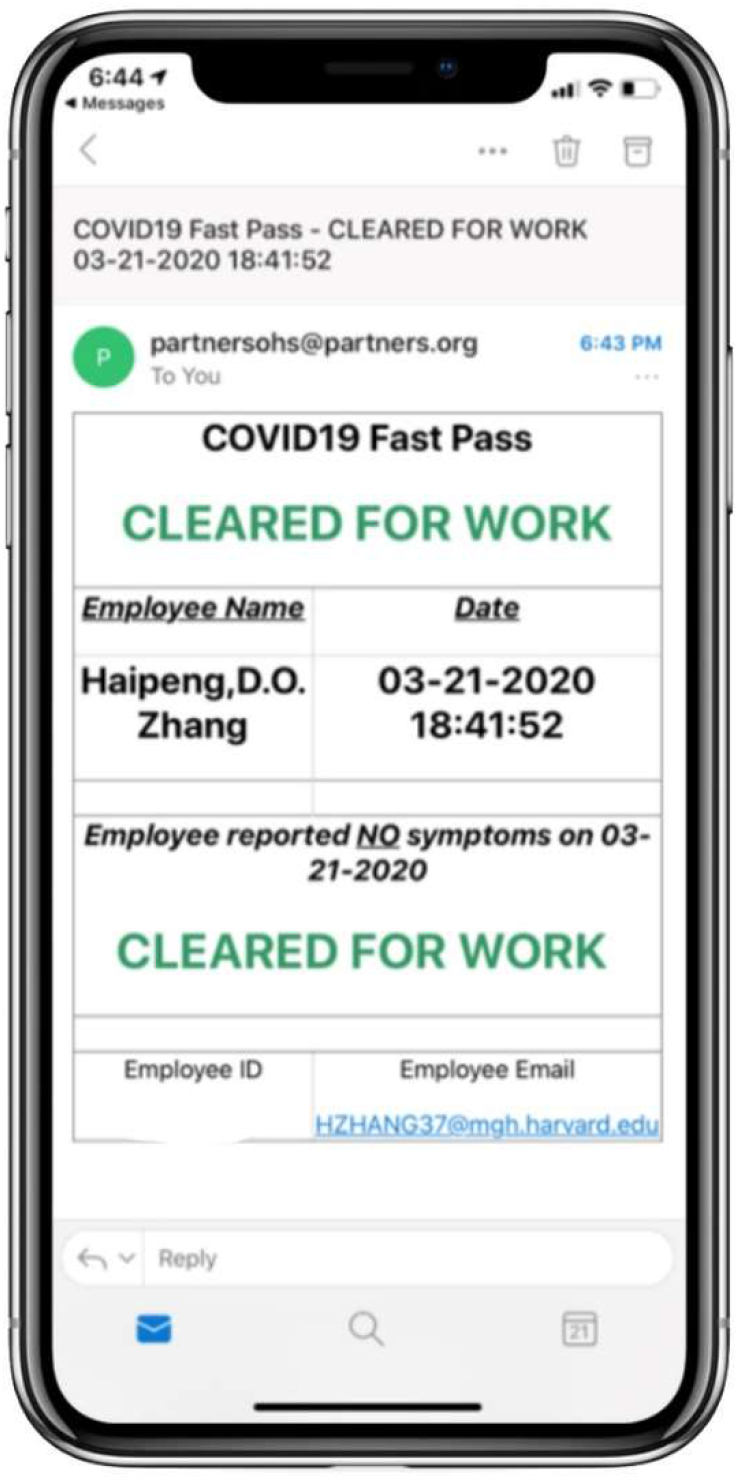
“Cleared for Work” version of COVID Pass.

**Figure 1C.**
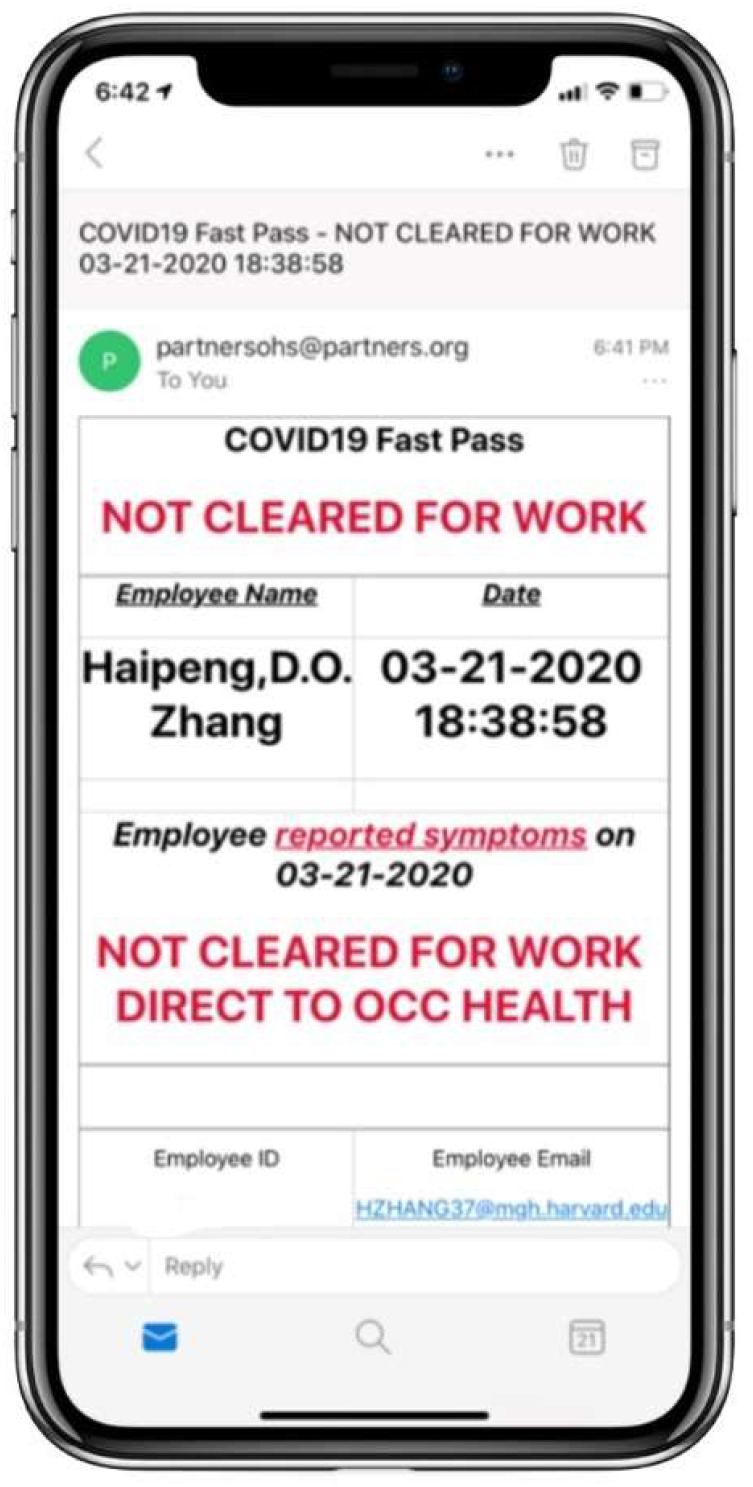
“Not Cleared for Work” version of COVID Pass.

We made COVID Pass available to employees via multiple methods. Working with marketing and internal communications experts, we selected an easy to remember Uniform Resource Locator (URL), https://www.partners.org/covidpass. We also created a Quick Response (QR) code for the URL so that users could use their smartphones to scan the QR code and be directed to the website. Finally, we wrapped the website in a native iOS and Android application so that the application could be distributed internally through our employee app stores.

For employees without access to a computer or mobile device to complete COVID Pass, there was a manual process established where in-person, socially distanced screeners asked employees entering the site if they have symptoms and then manually recorded the responses. For the manual pathway MVP, we created custom Microsoft Office 365 forms that allowed screeners at each of the pilot sites to quickly record the employees first name, last name, and whether they had symptoms.

A custom .NET Framework application was developed by a separate development team as the REDCap version of COVID Pass was being refined and requirements solidified. This version would include both the self-service attestation mode along with the manual process rolled into one build. As COVID Pass scaled, we ultimately transitioned from the REDCap application to the custom .NET application on March 30, 2020 at 5pm.

All versions of COVID Pass recorded employee attestations. The data were aggregated at the site level and shared with site operational leaders daily; individuals reporting positive symptoms were shared with Human Resources leadership daily. In this report, we share data from the first week of full enterprise COVID Pass use (March 30, 2020 – April 5, 2020) from the REDCap application database, the Office 365 spreadsheets, and the .NET Framework COVID Pass application. Aggregate descriptive statistics including overall employee attestations by day and site, employee attestations by application method (COVID Pass automatic screening vs. manual screening), employee attestations by time of day, and % of employees reporting COVID-19 symptoms were compiled using SAS (Enterprise Guide 7.1, SAS Institute Inc, Cary, NC) and Microsoft Excel (Microsoft Corporation, Redmond, WA). This study was approved by the Partners HealthCare Institutional Review Board.

The COVID Pass MVP was initially tested at two hospitals within our system on 3/23/20. As part of this controlled rollout, the project team participated in onsite testing of the application, gathering user feedback and being “at the elbow” with the staff at the entrance ways of sites. Figure 2 below shows an example hospital entrance with a COVID Pass lane for employees that completed COVID Pass prior to or immediately upon arrival. The manual screening lane includes a table for screeners to enter employee’s attestations manually. Feedback from both the staff assigned to screen employees and employees using COVID Pass were relayed to the developer for further refinement of the application. In parallel to this, development process, the number of sites within the system using COVID Pass slowly grew throughout the week. By 3/30/20, COVID Pass was deployed enterprise wide.

**Figure 2.**
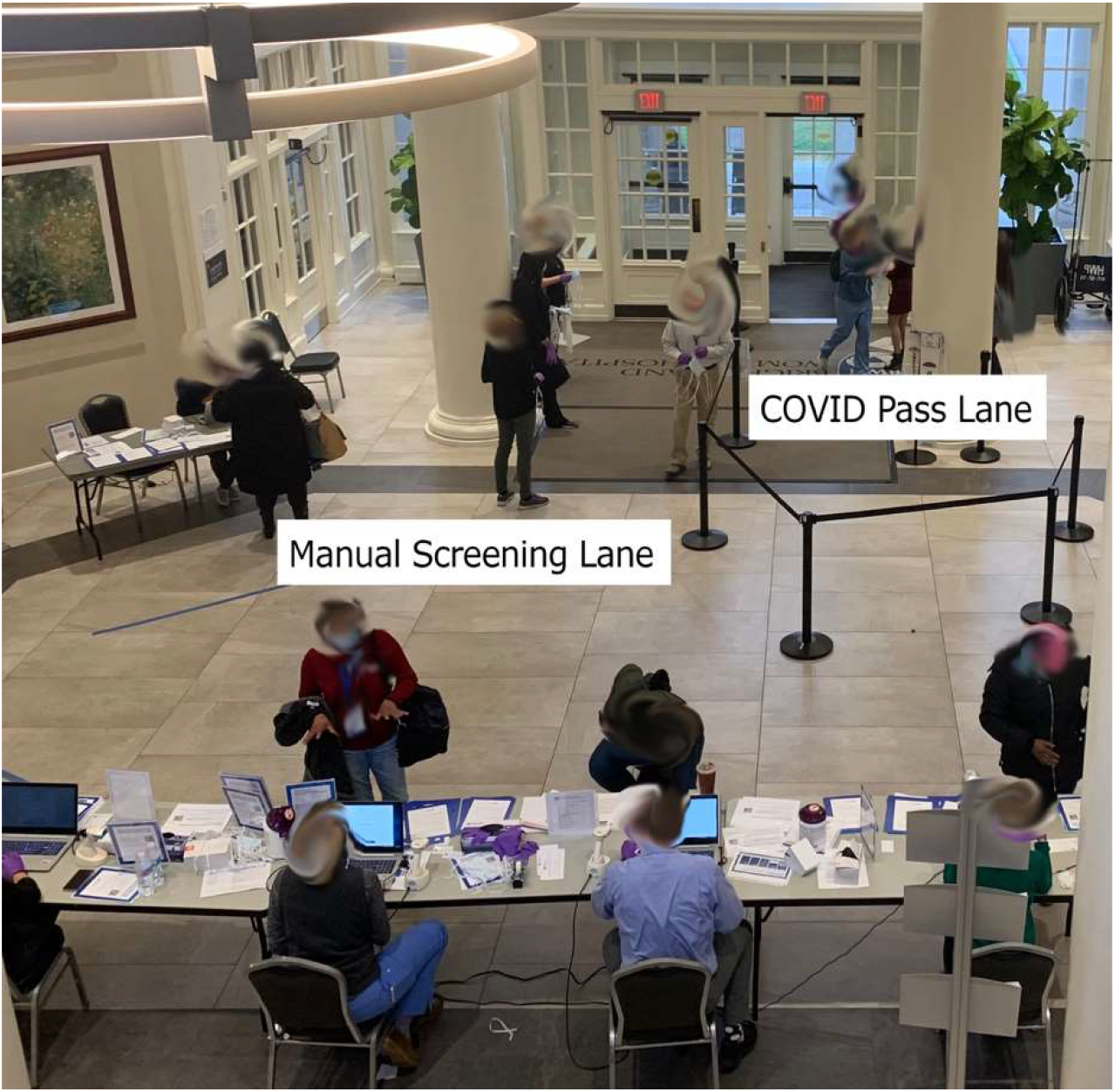
Hospital staff entrance showing COVID Pass lane for employees who used the COVID Pass application and the manual screening lane for employees who did not use the application

Documentation about initiating COVID Pass within a facility was collated and distributed to the sites as new COVID Pass sites went live. This documentation gave standard language, collateral such as flyers used in previous site implementations, and “lessons learned” from earlier implementations. The collective documentation also simplified the process of scaling this solution across the enterprise.

## Results

Over the course of the first full week of deployment across the enterprise (Mar 30, 2020 – April 5, 2020), 154,730 employee attestation logs were processed. Over this 7-day period, 558 (0.36%) employees reported positive symptoms. Table 1 summarizes the number of employee attestations (using either COVID Pass or manual process) by day and site. In most sites, most employees (∼80-90%) used the COVID Pass application, with a smaller percentage (∼10-20%) using manual attestation (Table 2). Hospital staff continued to work around the clock, but as expected staff attestations peaked during shift changes between 7-8am, 2-3pm, 4-6pm, and 11pm-midnight (Figure 3).

**Table 1.** Employee symptom attestations (COVID Pass + Manual) completions by day and site.

| SITE | COMPLETION_DATE |  |  |  |  |  |  | Total |
| --- | --- | --- | --- | --- | --- | --- | --- | --- |
|  | 30-Mar-20 | 31-Mar-20 | 1-Apr-20 | 2-Apr-20 | 3-Apr-20 | 4-Apr-20 | 5-Apr-20 |  |
| Site 1 | 8,676 | 8,251 | 8,318 | 8,316 | 7,901 | 3,986 | 3,788 | 49,236 |
| Site 2 | 7,190 | 7,140 | 7,144 | 7,047 | 6,691 | 3,012 | 2,804 | 41,028 |
| Site 3 | 2,268 | 2,142 | 2,152 | 2,151 | 2,113 | 1,030 | 1,008 | 12,864 |
| Site 4 | 2,578 | 2,148 | 1,874 | 1,952 | 1,778 | 876 | 842 | 12,048 |
| Site 5 | 1,376 | 1,372 | 1,376 | 1,372 | 1,295 | 462 | 438 | 7,691 |
| Site 6 | 916 | 1,208 | 1,214 | 1,200 | 1,167 | 533 | 514 | 6,752 |
| Site 7 | 876 | 828 | 826 | 821 | 824 | 338 | 348 | 4,861 |
| Site 8 | 654 | 677 | 767 | 819 | 792 | 362 | 380 | 4,451 |
| Site 9 | 613 | 458 | 748 | 708 | 643 | 319 | 266 | 3,755 |
| Site 10 | 571 | 634 | 632 | 682 | 627 | 100 | 90 | 3,336 |
| Site 11 | 136 | 254 | 401 | 514 | 520 | 109 | 78 | 2,012 |
| Site 12 | 430 | 398 | 406 | 395 | 368 | 69 | 63 | 2,129 |
| Site 13 | 15 | 275 | 321 | 378 | 372 | 194 | 160 | 1,715 |
| Site 14 | 8 | 188 | 232 | 201 | 192 | 117 | 126 | 1,064 |
| Site 15 | 11 | 209 | 189 | 203 | 205 | 81 | 58 | 956 |
| Site 16 | 637 | 2 | 2 | 1 | 1 | 0 | 0 | 643 |
| Site 17 | 1 | 20 | 11 | 13 | 5 | 2 | 24 | 76 |
| Site 18 | 0 | 9 | 13 | 16 | 15 | 3 | 3 | 59 |
| Site 19 | 6 | 9 | 16 | 10 | 4 | 7 | 2 | 54 |
| Site 20 | 0 | 0 | 0 | 0 | 0 | 0 | 0 | 0 |
| Total | 26,962 | 26,222 | 26,642 | 26,799 | 25,513 | 11,600 | 10,992 | 154,730 |

**Table 2.** COVID Pass employee self-attestation vs. Manual screening on April 2, 2020 by site. Note: some sites used separate manual screening data collection methods – their data are not reflected here.

| SITE | SOURCE |  |  |  |  |
| --- | --- | --- | --- | --- | --- |
|  | Electronic |  | Manual |  | Total |
|  | N | (%) | N | (%) |  |
| Site 1 | 8,316 | 100% | 0 | 0% | 8,316 |
| Site 2 | 6,288 | 89% | 759 | 11% | 7,047 |
| Site 3 | 1,883 | 88% | 268 | 12% | 2,151 |
| Site 4 | 1,711 | 88% | 241 | 12% | 1,952 |
| Site 5 | 1,372 | 100% | 0 | 0% | 1,372 |
| Site 6 | 973 | 81% | 227 | 19% | 1,200 |
| Site 7 | 819 | 100% | 2 | 0% | 821 |
| Site 8 | 744 | 91% | 75 | 9% | 819 |
| Site 9 | 364 | 51% | 344 | 49% | 708 |
| Site 10 | 632 | 93% | 50 | 7% | 682 |
| Site 11 | 514 | 100% | 0 | 0% | 514 |
| Site 12 | 395 | 100% | 0 | 0% | 395 |
| Site 13 | 295 | 78% | 83 | 22% | 378 |
| Site 14 | 161 | 80% | 40 | 20% | 201 |
| Site 15 | 203 | 100% | 0 | 0% | 203 |
| Site 16 | 1 | 100% | 0 | 0% | 1 |
| Site 17 | 13 | 100% | 0 | 0% | 13 |
| Site 18 | 16 | 100% | 0 | 0% | 16 |
| Site 19 | 10 | 100% | 0 | 0% | 10 |
| Site 20 | 0 | 0% | 0 | 0% | 0 |
| Total | 24,710 | 92% | 2,089 | 8% | 26,799 |

**Figure 3.**
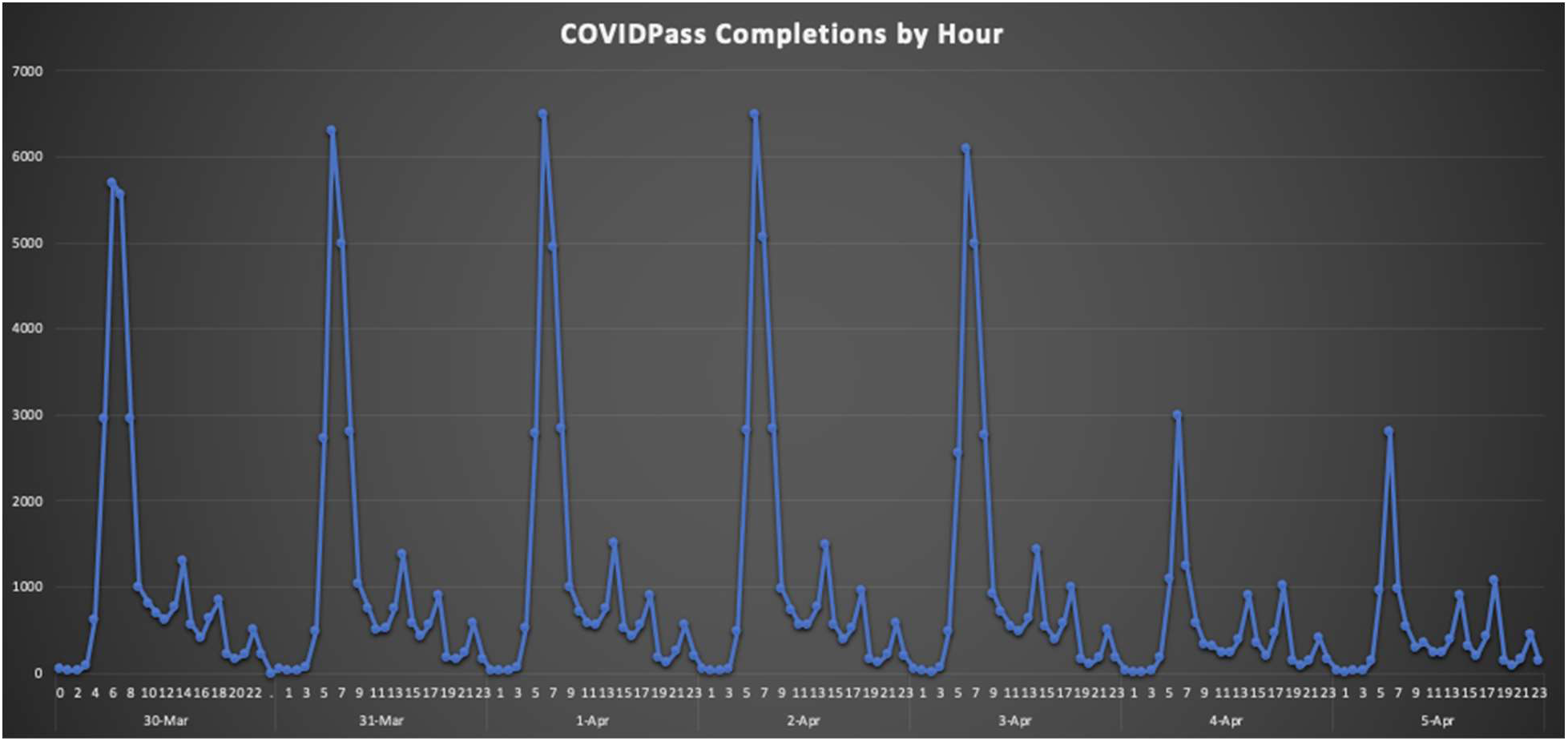
**Employee attestations (COVID Pass + Manual Screening) by hour of day over first week (March 30, 2020 – April 5, 2020)**

## Discussion

Within two weeks of conception, COVID Pass was live across the entire Partners HealthCare enterprise. During weekdays, COVID Pass recorded over 25,000 attestations a day. This high rate of adoption would not have occurred at such a fast pace without the following key considerations: 1) Mandatory and Incentivized Use while Minimizing Friction; 2) Planning for Accessibility; 3) Leveraging REDCap to enable an Agile development process; 4) Rapid analysis and distribution of COVID Pass data; and 6) Pre-emptive transition to more durable and scalable platform.

Making COVID Pass mandatory to gain access to work and pairing it with the distribution of masks quickly made the potential burden of a mandatory pre-work self-attestation more of a palatable proposition. COVID Pass would further benefit from the minimization of friction to complete the attestation process. We did this by minimizing the amount of interactions users would need to complete COVID Pass while maximizing the data captured. By auto-populating demographic information after login, the user only had to answer one question symptom review. If asymptomatic, we required attestation with their initials to receive the COVID Pass for the day. We also simplified access to COVID Pass through the creation of multiple distribution channels including a simple URL that was sent with all communication to staff throughout our organization, a QR code that was used on flyers in the entranceways of our facilities, and also an iOS and Android version of COVID Pass made available through our employee facing App Catalog.

As part of this work, we also knew that we would need to offer an accessible pathway for users who may not be able to complete the self-service COVID Pass due to limited proficiency with a smartphone or computer, language limitations, or other reasons. By incorporating this accessibility requirement early on in our development and creating a manual pathway, COVID Pass could then be a comprehensive solution for all employees.

Adopting an agile development process to further minimize friction for the end user through early on-site testing and rapid iteration cycles. An agile development approach emphasizes “early and continuous delivery of valuable software” while accommodating for changing requirements as software is utilized in the real world and assumptions are validated or invalidated. ^9^ Our team created the MVP for COVID Pass with 48 hours and began testing the solution the very next day. We refined the requirements for COVID Pass and made updates to the MVP multiple times throughout the day at the beginning of this process.

Much of this early ability to create such rapid changes and adjustments to COVID Pass came from the initial platform decision to utilize REDCap for the MVP. As the core functionality of REDCap was able to accommodate most of the initial requirements of COVID Pass, adjusting the symptom survey fields and conditional text could be done almost instantaneously. This enabled our team to sustain a rapid iteration cycle during the first week of deployment while additional sites across the organization went live with COVID Pass.

In addition, REDCap enabled our team to rapidly export data and custom reports from COVID Pass to our organization daily. This data was primarily utilized to do two major functions within the organization: 1) Tracking symptomatic employees for further workup of COVID-19 by occupational health; and 2) Estimate the number of employees on site for personal protective equipment supply planning.

Within the first week, COVID Pass had identified over 500 employees with symptoms that could be suggestive of COVID-19 infection. Further work is needed to correlate how many of these employees were actual confirmed cases. A preliminary estimate of R0 for COVID-19 is estimated to be somewhere between 2 and 4. ^10,11^ By identifying and instructing over 500 employees with symptoms suggestive of COVID-19 to not report to work, and alerting occupation health about these employees for further follow-up, COVID Pass has likely played an important role in limiting further spread of COVID-19 within our workforce, our patients, and the larger community in our region.

The World Health Organization estimates that up to 89 million medical masks are required monthly for the COVID-19 response. ^12^ As all COVID Pass logs who are cleared for work are issued a face mask, the daily logs from COVID Pass have become an important proxy to the number of masks being distributed daily to employees within our organization. This also gave leaders within the organization a good way to predict face mask allocations per facility, per day and plan accordingly.

By the middle of the first week of the COVID Pass rollout, it became apparent that a more hardened version of this application would be needed. Some of the clear advantages of REDCap such as the prebuilt scaffolding to support survey creation and data export became limitations as more custom change requests began to surface with COVID Pass. In addition, as COVID Pass rapidly become adopted enterprise wide, a more formalized support structure was needed to keep COVID Pass operational 24 hours a day, 7 days a week. As a result, we worked in parallel with a second, in-house development team to create a custom version of COVID Pass built on the Microsoft .NET Framework with a SQL database, utilizing the learnings and updated requirements gained through the release of the live REDCap application.

We ultimately transitioned to the .NET version of COVID Pass on March 30, 2020 at 5pm. The process of developing the same solution in parallel and in two different platforms allowed us to leverage the strengths of each one and achieve a more robust final product. The established processes and large user base for COVID Pass meant that a compiled, single-purpose application, such as the .NET application, was better suited for the needs of the project in the long-run. Transitioning the application to an infrastructure environment that provided 24/7 support and available resource capacity meant that the system could handle high load times during the morning hours when the highest number of concurrent users occurs. A refined self-service module and a manual pathway built on the same platform allowed for the streamlining of the data collection and analysis as well.

## Limitations

We custom developed COVID Pass to meet local requirements for employee symptom attestation. Other organizations may have different operational requirements. We are making both the REDCap and .NET source code available to other organizations to use and modify as needed. Secondly, some of our sites did not track manual attestation using the Office 365 forms or .NET applications, therefore we do not have complete data on manual screenings.

## Conclusion

Health systems across the world have faced incredible challenges from the COVID-19 pandemic. At Partners HealthCare, one challenge was the evolving role of symptom monitoring for all employees working onsite. Although technology is not a panacea, when used appropriately and scoped to the right problems, technology can play a meaningful role in the era of COVID-19.

With the combination of a rapid, agile approach to software development and a use case paired with an end-user incentive, we quickly created and deployed a dedicated employee attestation application that gained widespread adoption and use within our health system. COVID Pass is continuing to support daily screening for over 25,000 employees. Further, we have identified over 500 symptomatic employees that otherwise would have possibly come to work, potentially putting others at risk.

We continue to utilize the data that comes out of COVID Pass for incident planning, such as PPE supply. The COVID Pass platform also has the potential to be an effective communication tool to the workforce as well – one use case that we are exploring is utilizing the COVID Pass final page to inform employees of voluntary COVID-19 research studies within our institution.

As we share these lessons learned and the story of our implementation of COVID Pass, we also want to make sure that other institutions who may want to implement a similar solution can not only learn from our implementation but also have access to the source code for COVID Pass. Therefore, we are making the source code for COVID Pass available to all via GitHub at https://github.com/partnershealthcare.

These are extraordinary times we are all living through. We are confident that the bravery, commitment, and perseverance of the clinicians and hospital staff on the front lines of this crisis throughout the world will see us safely through COVID-19. Our hope is that software solutions like COVID Pass may play their small roles within this larger effort and that others may find meaningful use from this tool as our institution has.

## Data Availability

We have all data referred to in this manuscript and would happily share the de-identified data if needed.

